# Viral detection and identification in 20 minutes by rapid single-particle fluorescence in-situ hybridization of viral RNA

**DOI:** 10.1101/2021.06.24.21257174

**Authors:** Christof Hepp, Nicolas Shiaelis, Nicole C. Robb, Achillefs N Kapanidis

## Abstract

The increasing risk from viral outbreaks such as the ongoing COVID-19 pandemic exacerbates the need for rapid, affordable and sensitive methods for virus detection, identification and quantification; however, existing methods for detecting virus particles in biological samples usually depend on multistep protocols that take considerable time to yield a result. Here, we introduce a rapid fluorescence in situ hybridization (FISH) protocol capable of detecting influenza virus, avian infectious bronchitis virus and SARS-CoV-2 specifically and quantitatively in approximately 20 minutes, in both virus cultures and combined throat and nasal swabs without previous purification. This fast and facile workflow is applicable to a wide range of enveloped viruses and can be adapted both as a lab technique and a future diagnostic tool.

## Introduction

With over 150 million confirmed cases so far, COVID-19 is the most devastating worldwide pandemic since the Spanish Flu of 1918 [1-3]. As a respiratory infection caused by the coronavirus SARS-CoV-2, it first surfaced in China in late 2019 and has since resulted in over 3 million deaths worldwide. Contact tracing and rapid testing have been important tools to limit the spread of disease among the human population.

Currently, the most common diagnostic tests for a SARS-CoV-2 infection are based on nucleic acid amplification, antigen detection and serology tests [4]. Due to its high sensitivity and specificity, reverse transcriptase-mediated polymerase chain reaction (RT-PCR) represents the gold standard in SARS-CoV-2 testing. However, an RT-PCR test takes several hours to complete, and can only be performed in a laboratory setting, as it requires viral lysis and RNA purification procedures and therefore sample collection, transportation and reception. This routine commonly delays test results by 24 hours or more. As a promising alternative to RT-PCR, isothermal nucleic acid amplification methods such as loop-mediated isothermal amplification (LAMP) usually yield results within one hour [5-11]. However, both RT-PCR and LAMP testing have been compromised by supply chain issues due to expensive production and storage conditions of the required reagents. Rapid antigen-detection tests based on immunoassays and lateral flow formats have been developed to detect SARS-CoV-2, but these can suffer from low sensitivities [12]. Thus, alternative testing methods that yield quick results with low material costs are still urgently needed.

Fluorescence in situ hybridization (FISH) is a cytological technique that exploits the selective binding of fluorescently labeled oligo- and polynucleotides (20 – 20,000 nucleotides) to complementary sections of DNA and RNA inside fixed cells and tissues for the purpose of detection, quantification and spatial localization by a fluorescence microscope. In a medical context, FISH has predominantly been used to detect chromosomal aberrations and rearrangements in fixed cells, including translations, insertions and deletions [13-16]. FISH has been used to detect and identify pathogenic microbes, including bacteria [17, 18] and viral infections, e.g. HIV, Epstein-Barr virus and Dengue inside infected cells [19-22].

RNA FISH has been used extensively to visualize gene transcription in situ [23, 24]. By increasing the number of fluorophores that bind one RNA transcript, detection of single transcripts became possible, giving rise to single molecule FISH (smFISH) [25, 26]. Typical smFISH applications use an array of fluorescently labelled DNA oligonucleotides (probes) that bind to several sites on the viral genome, thereby accumulating several fluorescent dye molecules on a viral RNA, making it visible as a bright, diffraction-limited spot, while unspecific binding to the surface will lead to a darker background of single probes. Further, by utilizing smFISH probes that carry spectrally distinct fluorophores, more than one RNA species can be imaged simultaneously [27]. In virology, single viral genomes or gene segments can be detected by smFISH leading to new insights into the molecular mechanism of infection, replication and assembly of influenza viruses [28-30]. Similar studies have been performed in other viruses, e.g., rotavirus, bunyavirus and human leukaemia virus [31-33]. Importantly, isolated virus particles can also be also visualized by smFISH [34].

Using smFISH, single viral RNA molecules can be detected without the need for enzymatic amplification. However, typical FISH assay times still take several hours to an entire day [35]. By increasing the concentrations of FISH probes and optimizing fixation conditions, RNA FISH hybridization times could be reduced to 5 min [36]. Moreover, choosing target sequences that do not form secondary structures can greatly increase hybridization rates [37].

In this study, we introduce a rapid viral FISH protocol (rvFISH) for the detection of virus particles; the development of this improved assay depended on a systematic analysis of the efficiency of the hybridization reaction as we reduced the number of experimental steps and the hybridization time. We were able to detect influenza particles and infectious bronchitis virus (IBV), an avian coronavirus, down to a concentration of 10^5^ PFU/ mL and 10^2^ PFU/ mL, respectively, in a 20-minute assay. Further, we demonstrated that inactivated SARS-CoV-2 particles could be detected in the same way. Moreover, we showed that virus particles could be detected in nasopharyngeal swabs. By employing a dual-labeling strategy, we were able to suppress background caused by misidentified bright, fluorescent spots. Taken together, our data suggest that smFISH can be used as a powerful means to quickly and easily detect, identify and quantify a wide range of virus strains in both research and diagnostic applications.

## Results

### Development of a rapid viral FISH protocol for virus detection

As a starting point, we characterized the labelling efficiency of an existing RNA FISH protocol for staining viral RNA in the context of virus particles [35] [Fig 1A]. In this standard protocol, which takes ∼4 hours to complete, influenza virus particles were immobilised by evaporating the virus sample on a plasma-cleaned glass coverslip at room temperature. The virus particles were then fixed and the viral membrane was permeabilized to allow the probes to access the genome within. An array of fluorescently labelled DNA probes was hybridized to the specific target RNA, followed by several washing steps.

**Figure 1:**
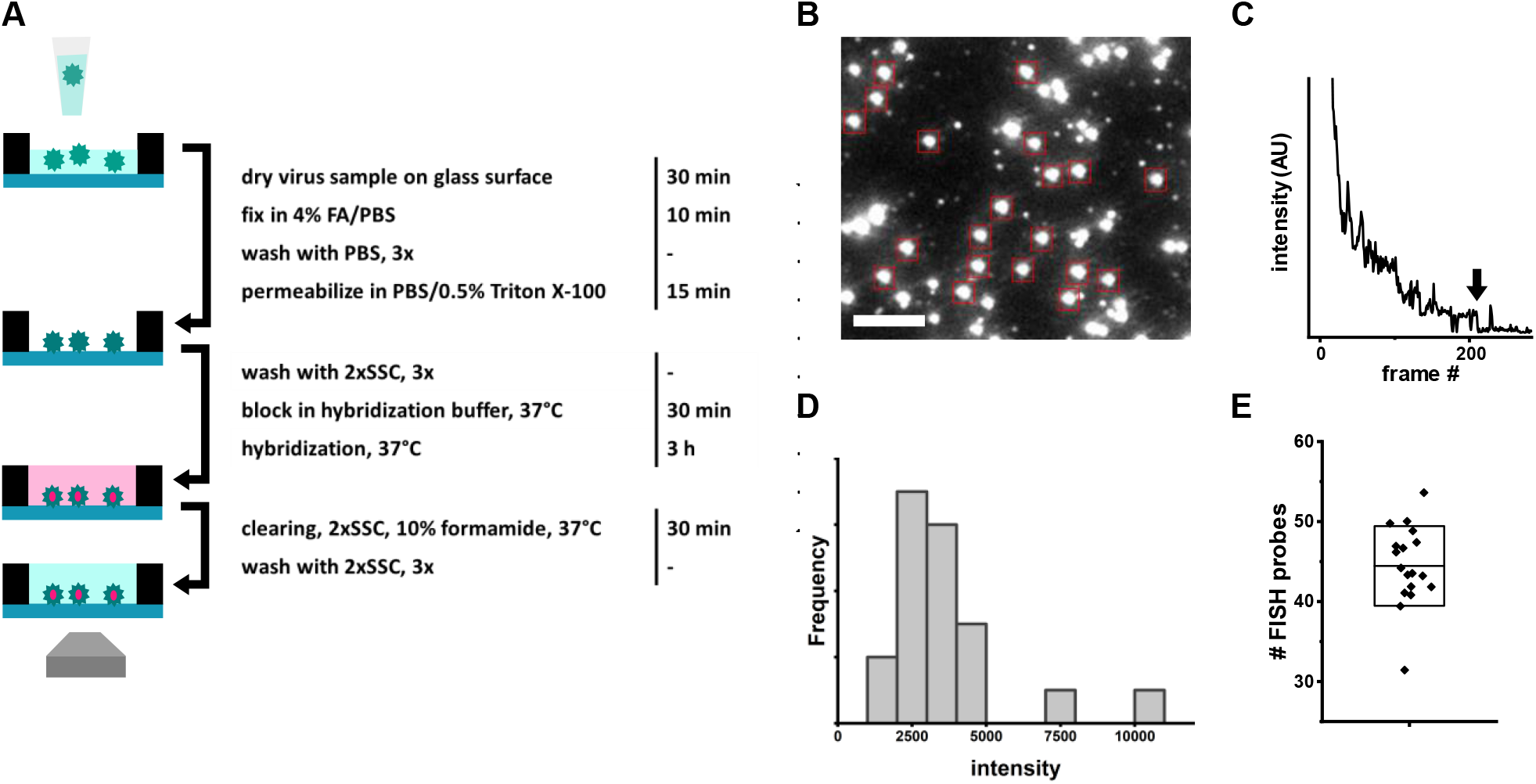
Efficiency of FISH in free virus particles. A) Schematic depiction and summary of the standard FISH based virus detection protocol with incubation times for the individual steps [35]. B) Representative field of view with influenza A/WSN/33 virus particles (10^6^ PFU/mL) stained by an array of 48 fluorescent hybridization probes. Red boxes: Diffraction limited, isolated particles that were analysed by stepwise photobleaching. Scale bar: 3 µm. C) Representative photobleaching curve recorded to determine the average number of probes bound to a virus particle. Arrow: Final bleaching step used to determine the average intensity of a single probe in a particle. D) Intensity distribution of the final bleaching steps of the particles in B (red boxes). E) Scatter plot of the probe counts for particles in B (red boxes). Box: average probe count per particle, standard deviation (SD).

To determine the labeling efficiency of this standard viral smFISH protocol, we calculated the number of probes bound to the neuraminidase (NA) gene of the influenza virus strain A/WSN/1933 (WSN). To achieve this, we immobilized the virus particles, stained them using a Quasar570-labelled FISH probe array comprising 48 individual 20-nucleotide (nt) probes, and used a single-molecule TIRF microscope to image the particles [Fig 1B] and count the bound FISH probes by stepwise photobleaching [Fig 1C]. By averaging the decrease in fluorescence intensity of the last bleaching step over several molecules, we determined the average fluorescence intensity corresponding to the single fluorophore of an individual FISH probe bound to a virus particle [Fig 1C, D]. The initial fluorescence intensities of the particles were then divided by the intensity of a single probe to yield the number of probes per particle [Fig 1D]. We found that the 48 targets on the NA segment were bound by an average of 45 FISH probes, translating to a very high binding efficiency (∼95%).

We then drastically shortened the protocol from ∼4h to 20 minutes [Fig. 2A] and evaluated the efficiency of the rapid viral FISH protocol. We optimized the conditions to a) achieve rapid and efficient immobilization of virus particles, b) reduce the hybridization time to a minimum and c) reduce the number of protocol steps. To enhance virus immobilization, the evaporation temperature was increased to 50 °C; at this temperature, a 20 µL sample took 10 min to evaporate, compared to 30 min in the standard protocol at room temperature. If the evaporation temperature was too high, e.g. 100 °C, the FISH signal was drastically reduced or absent. Evaporation was the most efficient method of immobilization, while interaction-based methods, e.g. PEG-Biotin, resulted in lower particle counts. Coating the glass surface with poly-L-lysine or chitosan did not increase the immobilization count, while occasionally resulting in a higher background from single probes. To reduce the hybridization time, we first considered that our results [Fig. 1] indicate that the hybridization conditions of the standard protocol result in a near-complete occupation of target sequences in isolated virus particles; this indicated that there should be conditions with decreased incubation time that should still provide high hybridization efficiency. Indeed, we found that after 10 min, ∼ 50% of the targets in a virus particle were occupied, enough to detect it with good accuracy. Regarding the reduction in the number of steps, we eliminated the blocking, fixation, clearing and most of the washing steps [Fig. 1A], as they did not substantially enhance the detection of isolated virus particles. However, it was necessary to wash once with 2x Saline Sodium Citrate (SSC) buffer after the immobilization to remove dried salts and media that interfered with FISH, as well as once with 2x SSC after hybridization to remove the bright fluorescent background of the probes in solution. The permeabilization and hybridization steps were combined by adding 0.25% Triton-X100 to the hybridization buffer.

**Figure 2:**
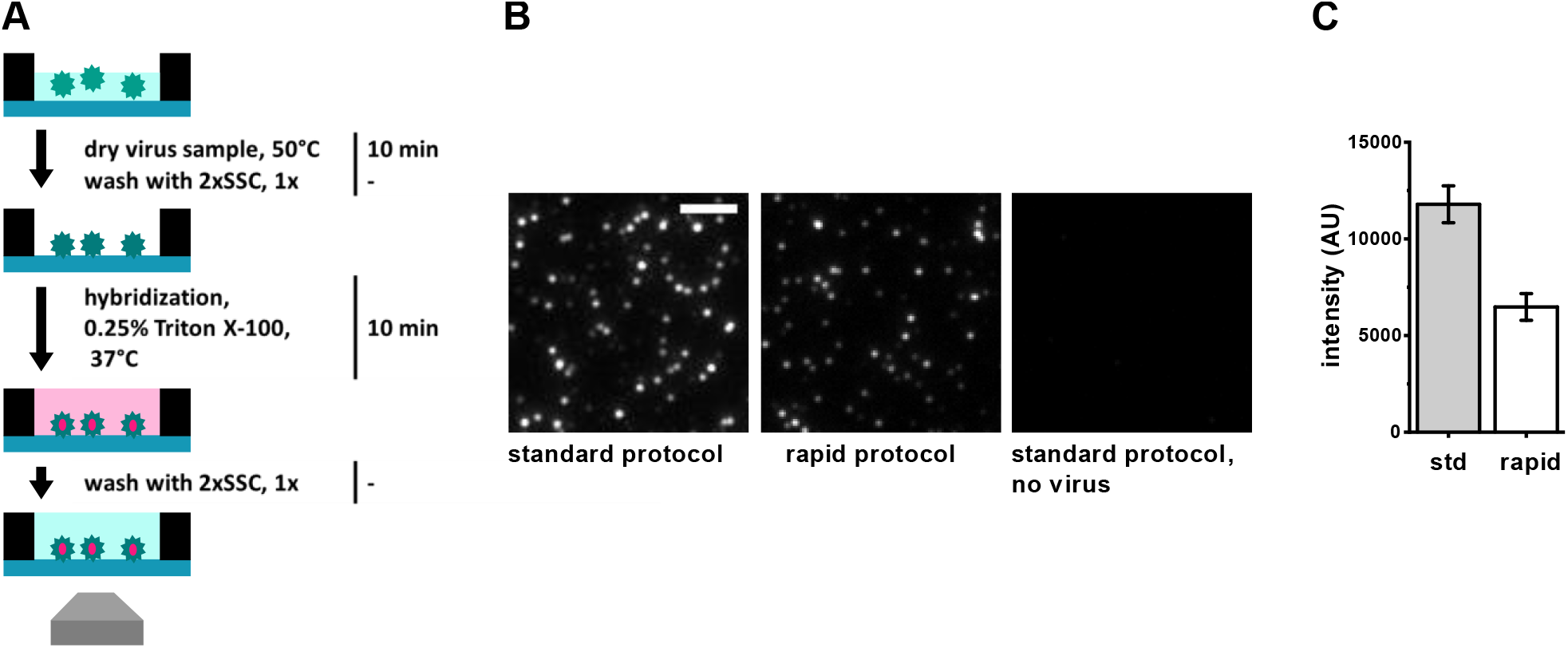
Efficiency of the rapid FISH protocol for virus detection. A) Schematic depiction and summary of the rapid virus detection FISH protocol with incubation times. B) Comparison of FISH protocols. Left panel: WSN particles stained by the standard protocol described in Fig. 1B [35], hybridization time 1h. Mid panel: WSN particles stained by the rvFISH protocol in Fig. 2A. Right panel: FISH staining (Fig. 1B) in the absence of virus. Scale bar: 3 µm. C) Particle intensity comparison of protocols in B. std: standard protocol, rapid: rapid protocol. Error bars: standard error of the mean (SEM).

Both the standard and rapid protocols gave rise to specific fluorescence signals on the surface, compared to a no-virus control [Fig. 2B], suggesting that the rvFISH protocol still allowed efficient staining of virus particles. By comparing the fluorescence intensities of the standard and rapid protocols, we found that the labelling efficiency of the rapid protocol was ∼50% of the standard protocol [Fig. 2C]. This suggests that particles are bound by ∼20 probes on average, making them easily distinguishable from single probes randomly bound to the surface.

### Detection of coronavirus using rapid RNA-FISH

Next, we determined whether the rvFISH assay could be used to detect coronavirus particles. We designed Quasar570-labelled probe arrays for the avian coronavirus (CoV) infectious bronchitis virus (IBV), comprising 32 individual 20-nt DNA probes. The probes targeted the nsp12 segment, encoding for RNA- dependent RNA polymerase (RdRP). We performed the rvFISH protocol on cultured influenza WSN strain (3.3 × 10^8^ plaque forming units (PFU)/mL) and IBV (10^6^ PFU/mL). For both stocks, experiments were performed and tested using the complementary FISH probe array (positive sample) and an array that targeted one of the other two stocks (negative control). We obtained bright signals for both virus strains when the complementary FISH probe sets were used; in contrast, we obtained only low or no background in the presence of the non-complementary probes [Fig. 3A], indicating that our labelling was specific.

**Figure 3:**
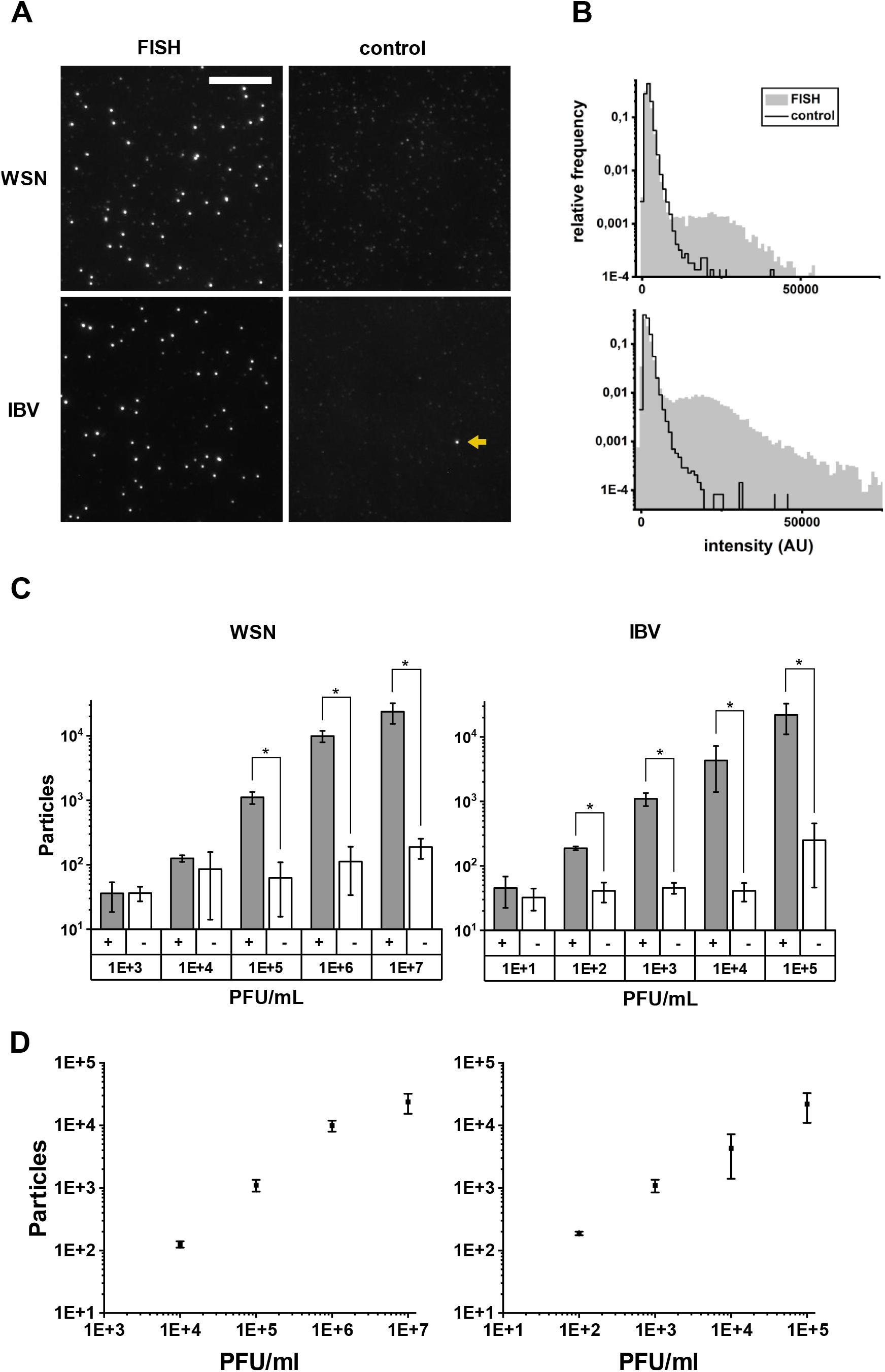
Performance of the rapid viral FISH protocol on virus culture supernatants of WSN and IBV in 0.9% NaCl. A) Images of FISH-stained WSN (top panel, 10^6^ PFU/mL) and IBV (mid panel, 10^4^ PFU/mL). Left column: correct FISH probe set, Right column: incorrect FISH probe set as a negative control: IBV for WSN, WSN for IBV. Arrow: Bright fluorescent spot in the negative sample that will be recognized as a virus particle. Scale bar: 10 µm. B) Intensity distributions of fluorescent spots detected in the samples in A. Top: WSN, bottom: IBV. Grey area: correct FISH probe set, black line: control FISH probe set (see A). C) Limit of detection (LOD) of FISH protocol for dilutions of virus culture supernatants in 0.9% NaCl. Left: Particle counts for an influenza sample. influenza probes (grey, +), IBV probes (white, +). Right: Particle counts for an IBV sample. IBV probes (grey, +), influenza probes (white, -), bottom panels: dilutions (PFU/mL), error bars: SD of three independent experiments, *T-test, p < 0.05. D) Particle counts with IBV probes plotted against PFU/mL. error bars: SD of three independent experiments.

We also generated distinct intensity distributions for the detected fluorescent spots in both positive sample and negative control data sets [Fig. 3B]. While negative samples showed a single distribution at low intensities, positive samples tended to result in bimodal distributions; these bimodal distributions included a low-intensity distribution (with a peak at ∼ 1,000 AU), which represents single probes and unspecific probe aggregates, and a high-intensity distribution (with a peak at ∼ 20,000 AU), which is caused by FISH-stained virus particles [Fig. 3B]. This was the case for both the WSN and IBV virus sample, illustrating that both WSN and IBV can be detected with the rvFISH assay.

### Determining the limit of detection for influenza and coronaviruses

Next, we determined the detection limit of the rvFISH assay from the fluorescence intensity distributions. To quantify the number of particles, we corrected their intensities for day-to-day fluctuations in laser power (about ± 20%) by normalizing them to the approximate intensity value of a single fluorophore, given by the median intensity value of the negative sample with the lowest virus concentration. Spots within an intensity range of 15-40 single fluorophores were counted as particles. While particles were present in both positive and negative samples, the number of detected particles was significantly higher (T-test, p < 0.05) in positive samples down to a concentration of 10^5^ PFU/ mL for WSN and 10^2^ PFU/ mL for IBV [Fig. 3C, D]. The detection sensitivity was limited by bright fluorescent spots occurring in the negative samples that were falsely identified as virus particles [Fig. 3A, top right, arrow]. Typically, a negative sample consisting of 81 FOVs had ∼ 100 such spots, and their number slightly increased with higher virus concentrations. These spots are most likely caused by FISH probe aggregates in the hybridization solution and several probe molecules non-specifically binding to distinct locations on the surface, and will subsequently be referred to as ‘noise’. The intensity distribution of bright spots in a negative sample is different from the virus samples in that the particle counts are continuously decreasing with higher intensities [Fig. 3B]. Overall, however, our limit of detection (LOD) was in a comparable range to that of other diagnostic tests, such as RT-PCR (LOD ∼10^2^ RNA copies/ mL) [38] or lateral flow assays (LOD ∼10^2^ PFU/ mL for SARS-CoV-2) [39].

### Detection of SARS-CoV-2 using rapid RNA-FISH

To demonstrate that the rvFISH assay was capable of detecting SARS-CoV-2, we designed a 44-probe FISH array targeting the nsp12 segment, analogous to the IBV probe set. The FISH protocol was performed on cultured SARS-CoV-2 (10^6^ PFU/ mL) that was diluted to 10^4^ PFU/ mL. Similar to WSN and IBV, a high number of bright particles was only seen in the presence of the SARS-CoV-2 probes [Fig. 4A]. Moreover, an IBV sample with SARS-CoV-2 probes had the same low number of bright spots as other negative samples, underlining the probe set specificity [Fig. 3A, bottom right]. The correct probe set resulted in a bimodal distribution, similar to WSN and IBV [Fig. 4B]. However, SARS-CoV-2 particles were generally not as bright as IBV particles, despite the higher number of probes in a SARS-CoV-2 FISH array (44 vs. 32), perhaps because SARS-CoV-2 was inactivated in 4% formaldehyde before use; formaldehyde has been shown to decrease the probe binding efficiency to mammalian cells in a previously established rapid FISH protocol [36]. Together with the LOD for IBV, this result suggests that the assay could be developed into a clinically relevant diagnostic test.

**Figure 4:**
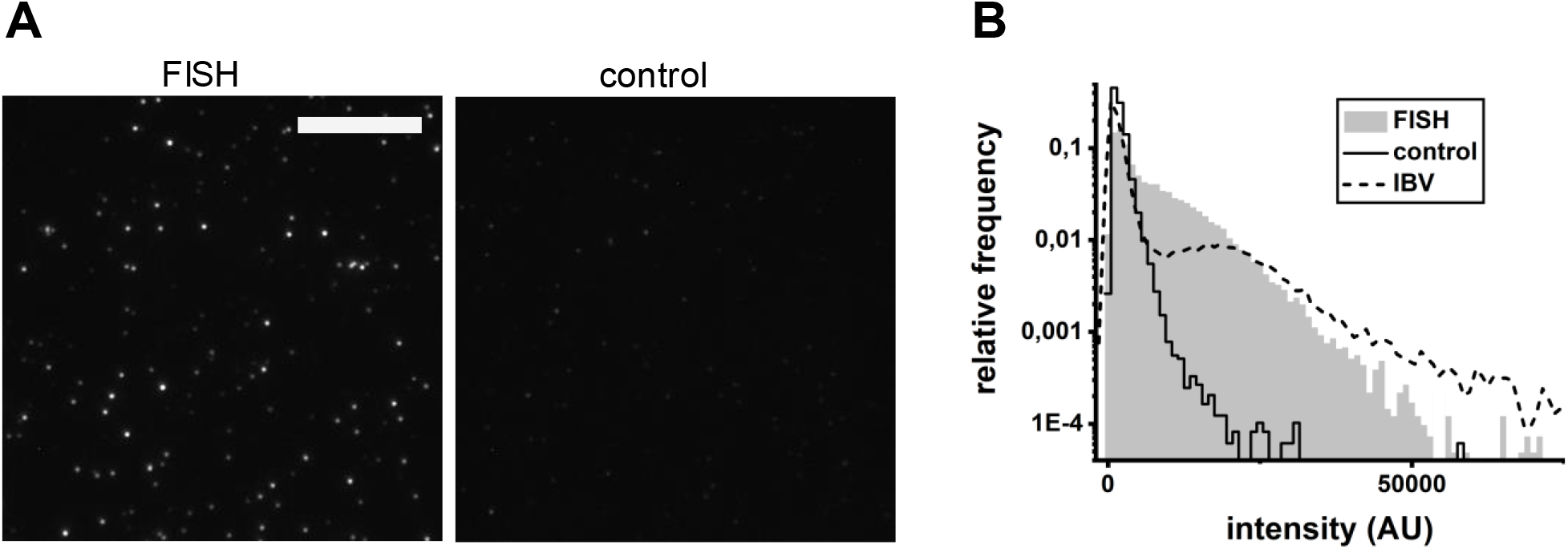
Performance of the virus detection FISH protocol on a virus culture supernatant of SARS-CoV- 2 in 0.9% NaCl. A) Images of FISH-stained SARS-CoV-2 (10^4^ PFU/mL). Left: SARS-CoV-2 FISH probe set, right: IBV probe set as a negative control. Scale bar: 10 µm. B) Intensity distributions of fluorescent spots detected in the samples in A. Grey area: complementary FISH probe set, black line: negative control FISH probe set, dotted line: IBV sample with IBV probes (Fig3B, bottom) for comparison.

### Dual-color rapid FISH to increase the assay specificity and suppress noise

As the detection sensitivity was limited by non-specific noise, we examined the possibility of noise reduction by staining WSN particles with two sets of WSN probes carrying spectrally distinct fluorophores. For this purpose, we split the WSN probe array described above into two separate arrays that were labeled with Quasar570 (green) and Quasar650 (red), respectively. The probes of both sets bind along the NA segment in an alternating manner [Fig. 5A]. In the detection assay, we incubated the sample with green probes only and subsequently added the red probes for another 5 min. This way, all unspecific binding sites should predominantly be occupied by the green probes, while the virus particles would be stained with both fluorescent dyes, resulting in co-localized spots in the green and the red channel [Fig. 5B]. As a negative control, we performed the assay on just the culture medium the viruses were propagated in (Dulbecco’s Minimal Essential Medium + 0.5% foetal calf serum), in the absence of virus particles. Spots in both the red and the green channel were analyzed as described above. However, as the total number of probes in the array was equal to that of the monochromatic probe set used earlier, 50% less probe binding was expected. Hence, spots with an intensity of 5-20 green fluorophores and 3-20 red fluorophores were counted as particles in the green and red channel, respectively. We found a signal-to-noise ratio (SNR) [Fig. 5C] similar to the single-color probe array [Fig. 4A]. However, when we only counted the particles that co-localized in the green and red channel, the noise was reduced 10-fold, while the particle counts in the positive sample were only reduced 1.6-fold [Fig. 5C]. A direct comparison of the SNR of the correlated spots from Fig. 5C to the performance of a monochromatic probe set of the same size [Fig. 4A] showed a 5-fold increase of the SNR when using dual-color probe set [Fig. 5D]. Together, these results suggest that the use of a dual-color probe set can efficiently decrease the LOD in our rapid detection assay by a factor of 5.

**Figure 5:**
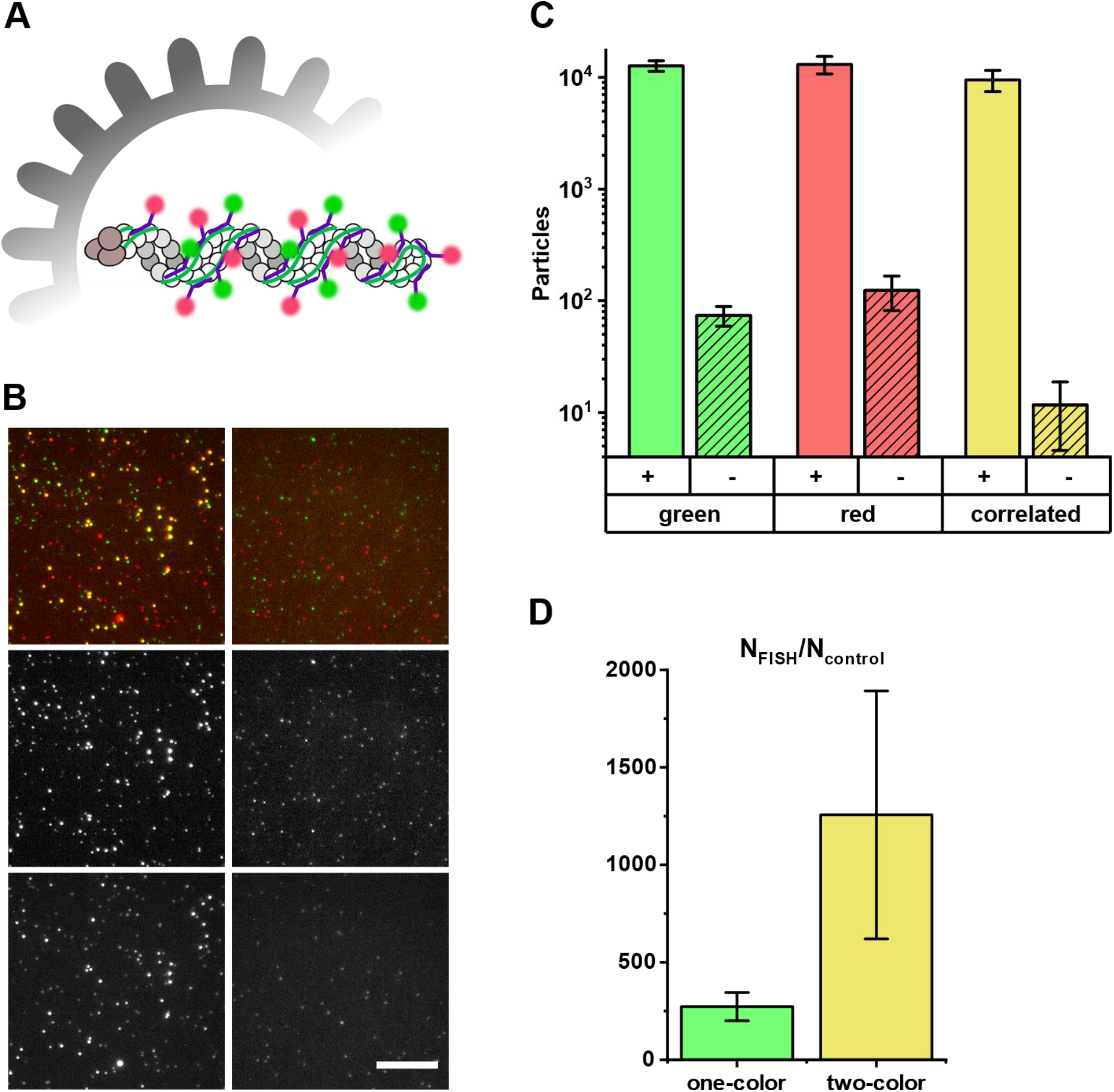
Increased signal-to-noise ratio with dual-color FISH. A) Schematic depiction of the NA segment in an influenza particle labelled with two probe sets carrying spectrally distinct fluorophores. B) Representative image of a dual-labelled influenza virus sample. Left column: diluted virus culture supernatant (10^6^ PFU/mL). Right column: equally diluted growth medium (negative control). Top row: composite image, middle row: green channel, bottom row: red channel. Scale bar: 10 µm. C) Particle counts in the green, red channel and co-localized particle-counts (yellow). Empty columns: virus sample, striped columns: negative control (DMEM + 5% FCS), error bars: SD, three independent experiments. D) The performance of a single-color FISH probe set (left) and the dual-color FISH probe set (right) with the same total number of probes is compared by the ratio of detected particles in the positive sample and the negative control. Error bars: SEM from three independent experiments.

### Performance of the FISH virus detection assay in combined nasal and throat swabs

In order to assess whether the rvFISH protocol would work on clinically relevant samples, we simulated virus detection in a combined throat and nasal swab. A swab was taken from an apparently healthy individual according to instructions published by Public Health England [Fig. 6A] [40]. To determine how the throat swab medium and any biological material from the patient affects the detection limit, we serially diluted the swab with 0.9% NaCl. To directly compare the particle counts, we added IBV to a final concentration of 10^4^ PFU/mL in all samples, a concentration that represents a compromise between a high number of detected virus particles and low noise. All samples were clarified by centrifugation and the rvFISH protocol was performed on them as described above. Additionally to IBV probes (Fig6B, “+”), we incubated the samples with SARS-CoV-2 probes as a negative control (“-“). In an undiluted throat swab, the particle count was drastically reduced and indistinguishable from the particle count of a negative sample [Fig. 6B]. For samples containing less throat swab material (diluted by 1:10, 1:100 and 1:1000), the particle count rose to up to ∼80% of the count obtained in the saline solution [Fig. 6B], indicating that diluting the throat swab material increases the overall sensitivity of the assay. However, as the dilution lowers the virus concentration, virus in throat swabs would best be detected by diluting the swabs as little as possible, as e.g., a 1:10 dilution would result in a detection efficiency of 0.1 × 23% = 2.3%, while a 1:100 dilution will only have an efficiency of 0.01 × 59 = 0.59%, etc. We conclude that the rvFISH assay will work on diluted clinical swab samples without purification, and suggest that material of non-viral origin in the swab, e.g., cellular debris and transfer medium ingredients, increases the LOD by about 1.5 to 2 orders of magnitude.

**Figure 6:**
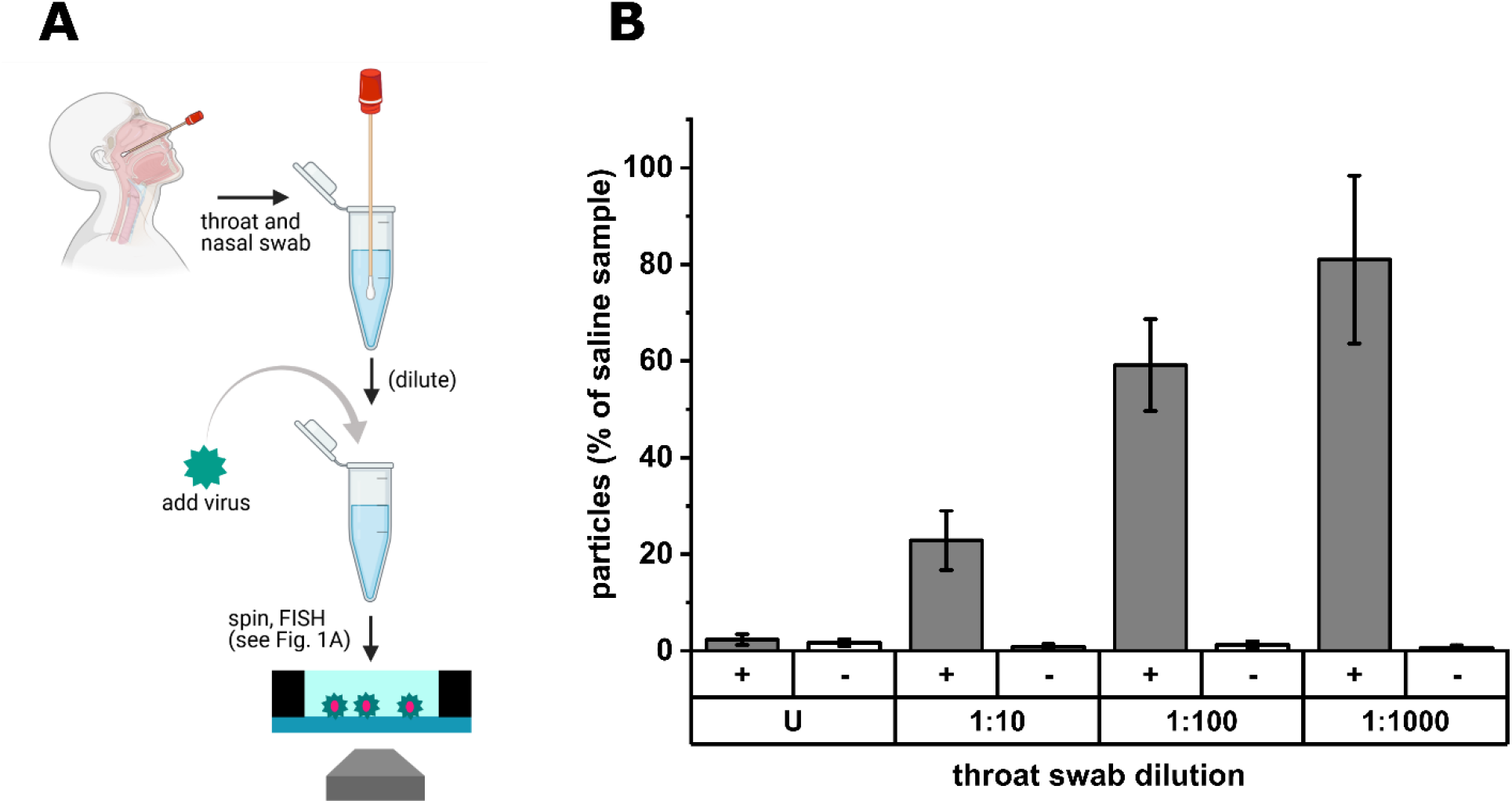
Efficiency of virus detection in a throat swab. A) Combined nasal and throat swabs were taken according to WHO instructions [40]. Subsequently, the throat swabs were diluted in saline and spiked with the same concentration of IBV (10^4^ PFU/mL) in each sample. After a centrifugation step, virus particles were immobilized and FISH stained as described. B) Efficiency of virus detection in throat swabs compared to virus diluted with saline, dependent on the dilution factor. [+]: IBV probes, [-]: SARS-Cov-2 probes, U: undiluted, error bars: SD of three independent experiments.

### Discussion

In this study, we explored the possibility of using a rapid FISH protocol for the detection of virus particles in culture supernatants and clinical samples. Using our protocol, we were able to detect influenza and coronavirus particles within 20 minutes after a sample is taken, substantially quicker than current RT-PCR protocols, while involving fewer protocol steps for sample purification [41]; our results open up the possibility of using FISH for real-time diagnosis and detection of virus infections. Moreover, our assay can be developed to rapidly measure virus titers in a virology lab, e.g. when biotechnological applications require a timely determination of the particle concentration in a virus culture. Direct comparison of the particle number determined by the FISH assay to the results from plaque assays can be used to determine the PFU-to-particle ratio, an important measure for the infectivity of a virus sample [42].

The current speed of detection is mainly limited by the evaporation time of the sample during immobilization and by the hybridization rate at the given probe concentration; to reduce the evaporation time of the virus sample, the temperature can be increased to above 50 °C. However, higher temperatures interfere with the FISH assay, through impacting either the stability of the viral RNA or the structure of the virus particles themselves, making them less likely to attach to the surface. Alternatively, a vacuum can be used to increase the evaporation rate, but might lead to increased technical requirements. A third solution involves passing the virus solution through a filter that retains virus particles and subsequently imaging the filter residue.

Regarding the hybridization time, previous rapid FISH protocols have demonstrated that probe concentrations of >100 µM can be used in eukaryotic cells, compared to 1 µM of probes used in our rapid smFISH assay [36]. Assuming that the hybridization rate increases linearly to probe concentration, we expect that the hybridization step can be further reduced to under one minute.

During the current COVID-19 pandemic, it became obvious that the need for SARS-CoV-2 testing exceeded the capacities of the biomedical sector and the biotech industry, and shortages occurred [43, 44]. FISH is less demanding in that it only uses fluorescent oligonucleotides and widely available lab chemicals that can be stored at room temperature, while other detection methods additionally require more expensive and sensitive biomolecules like enzymes, antibodies etc., making the rvFISH assay considerably cheaper than RT-PCR. Apart from *E. coli* tRNA, which can probably be replaced by a more durable blocking agent, all used materials can be stored at room temperature in dried form. The favourable storage conditions make our assay a good candidate for field use, e.g. in the context of emerging pathogens, and further exacerbate its capacity for real-time testing of viral infections.

The LOD was defined as the lowest tested virus concentration with statistically significant difference (t-Test, p<0.05) between a sample with a complementary probe set and a sample with a non-complementary control probe set. The LOD of our assay was 10^−5^ PFU/mL for WSN and 10^−2^ PFU/mL for IBV, comparable to the most sensitive lateral flow assays (LOD ∼10^2^ PFU/ mL for SARS-Cov-2) [39]. The discrepancy in LOD between the two virus strains can be explained by the observation that in general, not more than one in 10,000 cultured coronavirus particles is thought to be infective, while the particle-to-PFU ratio in influenza ranges from 10:1 to 100:1 [42, 45]. Therefore, qRT-PCR with a detection limit of 10^2^ RNA copies/ mL can still be considered at least 100 times more sensitive than our assay in its current form. However, not all RNA molecules in a virus sample might be accessible to PCR amplification, a scenario that is less likely for the binding of multiple FISH probes to an RNA target. To further lower the LOD of the rvFISH assay, the number of false-positive particles from fluorescent noise needs to be reduced, while the sampled area needs to be increased.

Regarding the reduction of false positives in the negative control, we note that, in our FISH data, we usually discovered ∼1 bright fluorescent spot per FOV that was identified as a virus particle by the detection algorithm, thereby limiting the sensitivity of our assay. We also demonstrated the success of a dual-colour labelling approach, which used co-localization analysis to reduce the number of false-positive particles 5-fold. The false positives can be reduced further by employing both a higher probe concentration and a larger probe array. The rationale in using more probes is that in a negative sample, false-positive particles become steadily less frequent at higher intensities; if virus particles have more probes bound to them, they will be brighter than and therefore better distinguishable from most of the noise. Further, in a previous study, we used deep learning to detect virus particles in culture supernatants and clinical samples by staining their surface with fluorescent oligonucleotides [46]. Here, deep learning may be a future option to recognize and eliminate false-positive particles from our FISH analysis.

Regarding the increase of sampled area, we used the difference in particle counts between positive and negative samples at an IBV concentration of 10^2^ PFU/ mL to estimate that we could observe ∼150 particles per dataset. This means that the theoretical detection limit in the absence of any noise would be around 10^0^ PFU/ mL, as the sampled area will contain less than one particle on average for lower concentrations. However, we only sampled an area of 0.33 mm^2^, compared to the total area of 28 mm^2^. Thus, sampling the entire area would increase the theoretical sensitivity by two orders of magnitude, comparable to RT-PCR. To achieve sampling of the total area in a reasonable time frame, the sampling velocity would have to be increased by using an objective with lower magnification, e.g., by using an objective with a magnification of 20x, instead of 100x, 25 times the area can be sampled in the same amount of time. However, depending on the power of the microscope, the use of weaker objectives might require enhancing the signal from the virus particles (see previous paragraph) due to their lower numerical aperture and light collection efficiency.

A practical limitation for the wide-spread application of our approach is the fact that a fluorescence microscope is required. In this study, we used a commercial instrument capable of detecting fluorescence from single molecules, that we used to calibrate the detection limit for virus particles. However, as brighter particles (like fluorescent beads) can be used for this calibration, single molecule detection is not a requirement, and virus particles (which bind ∼20 probes on average) should already be detectable by less sensitive microscopes. Improving particle brightness by expanding the probe arrays would bring down the technical requirements even more, to the point where mobile microscopy equipment might be used [47, 48]. The microscopy skills required from the testing personnel or experimenter can be reduced by automated acquisition, auto-focusing, or having a microscope with a fixed axial position. Apart from detecting a wide range of virus strains, the rvFISH protocol will be a versatile tool to design experiments to investigate hybridization of oligonucleotides to virus particles in detail and thus, provide a foundation for the optimization other hybridization-based techniques like PCR and LAMP.

## Methods

### Virus strains and FISH probe arrays

H1N1 A/WSN/1933 (WSN) influenza and coronavirus IBV have been described previously [46, 49]. Briefly, WSN virus was grown in Madin-Darby bovine kidney (MDBK) cells. The cell culture supernatant (Minimal Essential Media, Gibco) was collected and the viruses were titred by plaque assay, yielding a virus titre of 3.3 × 10^8^ plaque forming units (PFU)/mL. IBV (Beau-R strain)[50] was grown in chicken kidney (CK) cells and titred by plaque assay (1×10^6^ PFU/mL). Viruses were inactivated by addition of 0.2% formaldehyde before use. SARS-Cov-2 was grown in Vero E6 cells. The cell culture supernatant (Dulbecco’s Modified Eagle Medium, Gibco) was collected and titred by plaque assay, yielding a virus titre of 1.05×10^6^ PFU/mL. The virus was inactivated by addition of 4% formaldehyde before use.

Stellaris FISH probe arrays were ordered from Biosearch Technologies (Risskov, Denmark). The probes were designed using Stellaris’ online design tool, specifying the probe length as 20 nt and the maximum probe number per array as 48. The sequences of the NA segment in WSN and RdRP gene in CoV were selected as the target regions for the probes.

### Surface preparation

Coverslips (Menzel) were sonicated in 2% Hellmanex for 15 min, rinsed with MilliQ water 5 times, sonicated in water for another 5 min, rinsed with water 5 times and dried under an N2 stream. Subsequently, the dried coverslips were cleaned for 30 min in a plasma cleaner (Harrick) at high plasma strength for at least 30 min. Silicone gaskets (CultureWell, diameter 6 mm) were placed on the plasma-treated coverslips.

### Standard FISH protocol

This protocol is a modified version of a previously described protocol [35]. A virus stock diluted in 0.5% NaCl was added to a plasma-treated coverslip and evaporated at room temperature and washed with phosphate buffered saline (PBS). The immobilized virus was fixed with 4% formaldehyde in PBS for 10 min and subsequently permeabilized with 0.5% Triton X-100 in PBS for 15 min. After washing with 2x saline sodium citrate (SSC), the surface was blocked with hybridization buffer 1 (HB1; 2x SSC, 10% formamide, 10% dextran sulfate, 0.02% RNase free-BSA, 0.2 mg/ml E. coli tRNA, 1% RNasin Plus (Promega)) for 30 min at 37°C. Subsequently, the sample was incubated with 1 µM FISH probes in HB1 for 1-3h at 37°C. After hybridization, the sample was cleared with 2x SSC, 10% formamide, 1 % RNasin Plus and washed three times with 2x SSC.

### Rapid FISH protocol

A virus stock diluted in 0.5% NaCl was added to a plasma-treated coverslip and evaporated at 50°C for 10 min and washed with 2x SSC. The immobilized virus was incubated with 1 µM FISH probes in HB2 (2x SSC, 10% formamide, 0.02% RNase free-BSA, 0.2 mg/ml E. coli tRNA, 1% RNasin Plus, 0.25% Triton X-100) for 10 min at 37°C and washed once with 2x SSC after hybridization.

### Throat swabs

A swab was taken from an apparently healthy individual according to instructions published by Public Health England [40]. Experiments using throat swab material were performed in accordance with the Declaration of Helsinki and approved by the Medical Sciences Interdivisional Research Ethics Committee of the University of Oxford (R76273/RE001). Informed consent was obtained from the throat swab donor.

### Imaging conditions

All experiments were performed on a commercially available Nanoimager fluorescence microscope (Oxford Nanoimaging). The sample was imaged using total internal reflection fluorescence (TIRF) microscopy. The laser illumination was focused at an angle of 54° with respect to the default position. Images of a field of view (FOV) measuring 80 × 49 µm were taken with an exposure time of 500 ms, with a laser intensity kept constant at 780 kW/cm^2^ for the green (532 nm) laser when taking single-colour images, and at 624 kW/cm^2^ for the green laser and 390 kW/cm^2^ for the red (647 nm) laser, when taking dual-colour images. To automate the task and ensure no bias in the selection of FOVs, the whole sample was scanned using the multiple acquisition capability of the microscope; 81 FOVs were imaged in 3 minutes. For photobleaching experiments, the exposure time was decreased to 100 ms and the laser power (532 nm) was increased to 2.34 mW/cm^2^.

### Data analysis

All microscope data was processed using MatLab2019b. To correct the images for uneven illumination, an image was generated from the median pixel values across all FOVs and fitted with a cubic smoothing spline (‘csaps’, smoothing parameter = 10^−7^). The values of the fit were then normalized to the average intensity of a pixel and multiplied with the grey values of each raw image. Moreover, the average value of the fit was used as the fluorescent background of the images.

To detect fluorescent spots, each image was bandpass filtered (‘bpass’, noise length = 0, object length = 6). The median value of all non-zero pixels in the filtered image was multiplied by three and used as a threshold to binarize the filtered image. The ‘bwpropfilt’ function was used to exclude objects smaller than 5 pixels (px). The intensity-weighted centroids of the remaining regions were calculated from the grey values of the original image using the ‘regionprops’ function (Steve Eddins). The intensity of the fluorescent spot was determined as the background-subtracted intensity of a bounding box with a diameter of 5 px around the centroid.

## Data Availability

All data will be available upon request.

## Acknowledgements

Special thanks to Dr Erica Bickerton (Pirbright Institute, UK) for providing IBV, and Dr Rebecca Moore and Prof William James (Sir William Dunn School of Pathology, Oxford, UK) for providing SARS-CoV-2. Moreover, we thank the lab of Prof Ervin Fodor (Sir William Dunn School of Pathology, Oxford, UK) for helpful discussions, as well as Dr Leon Peto, Dr Nicole Stoesser and Prof Derrick Crook (Nuffield Department of Medicine, Oxford) for experimental support. This research was supported by a Royal Society Dorothy Hodgkin Research Fellowship DKR00620 (N.C.R.), the University of Oxford COVID-19 Research Response Fund (N.C.R and A.N.K.), a BBSRC-funded studentship (N.S.), BBSRC grant BB/V001868/1 (C.H., N.C.R. and A.N.K.) and Wellcome Trust grant 110164/Z/15/Z (A.N.K.). All data will be available upon request.

## Author Contributions

C.H. carried out experiments and analysed results. C.H., N.C.R and A.N.K. designed experiments and interpreted results. C.H. and N.S. wrote analysis software. C.H. wrote the manuscript; A.N.K and N.R. edited the manuscript. All authors reviewed and approved the final manuscript.

## References

1. Chan, J.F., et al., A familial cluster of pneumonia associated with the 2019 novel coronavirus indicating person-to-person transmission: a study of a family cluster. Lancet, 2020. 395(10223): p. 514–523.

2. Taubenberger, J.K. and D.M. Morens, 1918 Influenza: the mother of all pandemics. Emerg Infect Dis, 2006. 12(1): p. 15–22.

3. Hu, B., et al., Characteristics of SARS-CoV-2 and COVID-19. Nat Rev Microbiol, 2021. 19(3): p. 141–154.

4. Udugama, B., et al., Diagnosing COVID-19: The Disease and Tools for Detection. ACS Nano, 2020. 14(4): p. 3822–3835.

5. Huang, W.E., et al., RT-LAMP for rapid diagnosis of coronavirus SARS-CoV-2. Microb Biotechnol, 2020. 13(4): p. 950–961.

6. Lu, R., et al., Development of a Novel Reverse Transcription Loop-Mediated Isothermal Amplification Method for Rapid Detection of SARS-CoV-2. Virol Sin, 2020. 35(3): p. 344–347.

7. Lu, R., et al., Correction to: Development of a Novel Reverse Transcription Loop-Mediated Isothermal Amplification Method for Rapid Detection of SARS-CoV-2. Virol Sin, 2020. 35(4): p. 499.

8. El-Tholoth, M., H.H. Bau, and J. Song, A Single and Two-Stage, Closed-Tube, Molecular Test for the 2019 Novel Coronavirus (COVID-19) at Home, Clinic, and Points of Entry. ChemRxiv, 2020.

9. Lamb, L.E., et al., Rapid detection of novel coronavirus/Severe Acute Respiratory Syndrome Coronavirus 2 (SARS-CoV-2) by reverse transcription-loop-mediated isothermal amplification. PLoS One, 2020. 15(6): p. e0234682.

10. Zhang, Y., et al., Rapid Molecular Detection of SARS-CoV-2 (COVID-19) Virus RNA Using Colorimetric LAMP. 2020: p. 2020.02.26.20028373.

11. Rodriguez-Manzano, J., et al., A handheld point-of-care system for rapid detection of SARS-CoV-2 in under 20 minutes. 2020: p. 2020.06.29.20142349.

12. Chartrand, C. and M. Pai, How accurate are rapid influenza diagnostic tests? Expert Rev Anti Infect Ther, 2012. 10(6): p. 615–7.

13. Carter, N.P., et al., Reverse chromosome painting: a method for the rapid analysis of aberrant chromosomes in clinical cytogenetics. J Med Genet, 1992. 29(5): p. 299–307.

14. Weimer, J., M. Kiechle, and N. Arnold, FISH-microdissection (FISH-MD) analysis of complex chromosome rearrangements. Cytogenet Cell Genet, 2000. 88(1-2): p. 114–8.

15. Cherif, D., O. Bernard, and R. Berger, Detection of single-copy genes by nonisotopic in situ hybridization on human chromosomes. Hum Genet, 1989. 81(4): p. 358–62.

16. Bishop, R., Applications of fluorescence in situ hybridization (FISH) in detecting genetic aberrations of medical significance. Bioscience Horizons: The International Journal of Student Research, 2010. 3(1): p. 85–95.

17. Fazli, M., et al., PNA-based fluorescence in situ hybridization for identification of bacteria in clinical samples. Methods Mol Biol, 2014. 1211: p. 261–71.

18. Batani, G., et al., Fluorescence in situ hybridization (FISH) and cell sorting of living bacteria. Sci Rep, 2019. 9(1): p. 18618.

19. Hart, L., et al., Detection of human immunodeficiency virus in infected CEM cells using fluorescent DNA probes and a laser-based computerized image cytofluorometry system. Anal Quant Cytol Histol, 1990. 12(2): p. 127–34.

20. Chen, H.L., et al., Detection of EBV in nasopharyngeal carcinoma by quantum dot fluorescent in situ hybridization. Exp Mol Pathol, 2010. 89(3): p. 367–71.

21. Raquin, V., et al., Detection of dengue group viruses by fluorescence in situ hybridization. Parasit Vectors, 2012. 5: p. 243.

22. Frickmann, H., et al., Fluorescence in situ hybridization (FISH) in the microbiological diagnostic routine laboratory: a review. Crit Rev Microbiol, 2017. 43(3): p. 263–293.

23. Singer, R.H. and D.C. Ward, Actin gene expression visualized in chicken muscle tissue culture by using in situ hybridization with a biotinated nucleotide analog. Proc Natl Acad Sci U S A, 1982. 79(23): p. 7331–5.

24. Young, A.P., D.J. Jackson, and R.C. Wyeth, A technical review and guide to RNA fluorescence in situ hybridization. PeerJ, 2020. 8: p. e8806.

25. Femino, A.M., et al., Visualization of single RNA transcripts in situ. Science, 1998. 280(5363): p. 585–90.

26. Raj, A., et al., Imaging individual mRNA molecules using multiple singly labeled probes. Nat Methods, 2008. 5(10): p. 877–9.

27. Levsky, J.M., et al., Single-cell gene expression profiling. Science, 2002. 297(5582): p. 836–40.

28. Chou, Y.Y., et al., Colocalization of different influenza viral RNA segments in the cytoplasm before viral budding as shown by single-molecule sensitivity FISH analysis. PLoS Pathog, 2013. 9(5): p. e1003358.

29. Lakdawala, S.S., et al., Influenza a virus assembly intermediates fuse in the cytoplasm. PLoS Pathog, 2014. 10(3): p. e1003971.

30. Yamauchi, Y., Quantum dots crack the influenza uncoating puzzle. Proc Natl Acad Sci U S A, 2019. 116(7): p. 2404–2406.

31. Periz, J., et al., Rotavirus mRNAS are released by transcript-specific channels in the double-layered viral capsid. Proc Natl Acad Sci U S A, 2013. 110(29): p. 12042–7.

32. Erick, B.-M., et al., Research Square, 2021.

33. Billman, M.R., D. Rueda, and C.R.M. Bangham, Single-cell heterogeneity and cell-cycle-related viral gene bursts in the human leukaemia virus HTLV-1. Wellcome Open Res, 2017. 2: p. 87.

34. Chou, Y.Y., et al., One influenza virus particle packages eight unique viral RNAs as shown by FISH analysis. Proc Natl Acad Sci U S A, 2012. 109(23): p. 9101–6.

35. Chou, Y.Y. and T. Lionnet, Single-Molecule Sensitivity RNA FISH Analysis of Influenza Virus Genome Trafficking. Methods Mol Biol, 2018. 1836: p. 195–211.

36. Shaffer, S.M., et al., Turbo FISH: a method for rapid single molecule RNA FISH. PLoS One, 2013. 8(9): p. e75120.

37. Zhang, Z., et al., Single-molecule tracking of the transcription cycle by sub-second RNA detection. Elife, 2014. 3: p. e01775.

38. Arnaout, R., et al., SARS-CoV2 Testing: The Limit of Detection Matters. 2020: p. 2020.06.02.131144.

39. Peto, T., COVID-19: Rapid Antigen detection for SARS-CoV-2 by lateral flow assay: a national systematic evaluation for mass-testing. 2021: p. 2021.01.13.21249563.

40. How to use the self-swabbing kit for a combined throat and nose swab (video) 2020; Available from: https://www.gov.uk/government/publications/covid-19-guidance-for-taking-swab-samples/how-to-use-the-self-swabbing-kit-for-a-combined-throat-and-nose-swab-video.

41. Vogels, C.B.F., et al., Analytical sensitivity and efficiency comparisons of SARS-CoV-2 RT-qPCR primer-probe sets. Nat Microbiol, 2020. 5(10): p. 1299–1305.

42. Fonville, J.M., et al., Influenza Virus Reassortment Is Enhanced by Semi-infectious Particles but Can Be Suppressed by Defective Interfering Particles. PLoS Pathog, 2015. 11(10): p. e1005204.

43. The Scientist Mar 11, 2020; Available from: https://www.the-scientist.com/news-opinion/rna-extraction-kits-for-covid-19-tests-are-in-short-supply-in-us-67250.

44. Fomsgaard, A.S. and M.W. Rosenstierne, An alternative workflow for molecular detection of SARS-CoV-2 - escape from the NA extraction kit-shortage, Copenhagen, Denmark, March 2020. Euro Surveill, 2020. 25(14).

45. Klimstra, W.B., et al., SARS-CoV-2 growth, furin-cleavage-site adaptation and neutralization using serum from acutely infected hospitalized COVID-19 patients. J Gen Virol, 2020. 101(11): p. 1156–1169.

46. Shiaelis, N., et al., Virus detection and identification in minutes using single-particle imaging and deep learning. 2020: p. 2020.10.13.20212035.

47. Diederich, B., et al., A versatile and customizable low-cost 3D-printed open standard for microscopic imaging. Nat Commun, 2020. 11(1): p. 5979.

48. Trofymchuk, K., et al., Addressable nanoantennas with cleared hotspots for single-molecule detection on a portable smartphone microscope. Nat Commun, 2021. 12(1): p. 950.

49. Robb, N.C., et al., Rapid functionalisation and detection of viruses via a novel Ca(2+)-mediated virus-DNA interaction. Sci Rep, 2019. 9(1): p. 16219.

50. Casais, R., et al., Reverse genetics system for the avian coronavirus infectious bronchitis virus. J Virol, 2001. 75(24): p. 12359–69.

